# Myocardial performance improvement after iron replacement in heart failure patients: The IRON-PATH II echo-substudy

**DOI:** 10.1101/2025.02.19.25322580

**Authors:** Raúl Ramos-Polo, Maria del Mar Ras-Jiménez, María del Carmen Basalo Carbajales, Sílvia Jovells-Vaqué, José Manuel Garcia-Pinilla, Marta Cobo-Marcos, Javier de Juan-Bagudá, Cândida Fonseca, Josep Francesch Manzano, Andreea Eunice Cosa, Sergi Yun-Viladomat, Cristina Enjuanes, Marta Tajes Orduña, Josep Comin-Colet

## Abstract

Iron deficiency (ID) is a prevalent comorbidity in heart failure (HF) patients associated with poor prognosis and impaired physical capacity. Functional limitations linked to ID may be related to cardiac function abnormalities, which could be reversible with iron repletion. Some echocardiographic parameters, such as global longitudinal strain (GLS), myocardial work (MW), and its derivatives constructive work (CW), wasted work (WW), and work efficiency (WE), may provide additive value in advanced cardiac performance assessment. The IRON-PATH II was a multicenter, prospective and observational study designed to describe pathophysiological pathways associated to ID. The echo-substudy included 100 HF patients undergoing a specific pilot echocardiographic evaluation. Patients had left ventricular ejection fraction (LVEF) ≤50%, were in stable clinical condition, on standard HF medication, and with hemoglobin ≥11 g/dL. The final cohort included 98 patients. The ID group showed worse cardiac function, with lower GLS (–8.5±9% vs –10±10%), WE (74±10% vs 80±10%), and MW (665[453-1013] vs 947[542-1199] mmHg%), as well as higher WW (290[228-384] vs 212[138-305] mmHg%) and lower RV free wall strain (–13[-20-(–11)] % vs –17[-23-(–14)] %). Following iron repletion, ID patients demonstrated improved LV (GLS, MW, WE, and WW) and RV performance (RV free wall strain), aligning with non-ID patients (all p-values >0.05 compared to the non-ID group). In conclusion, among HF patients with reduced LVEF, ID was associated with worse myocardial performance in both the LV and RV, with these alterations being reversible after intravenous iron repletion.

Graphical Abstract

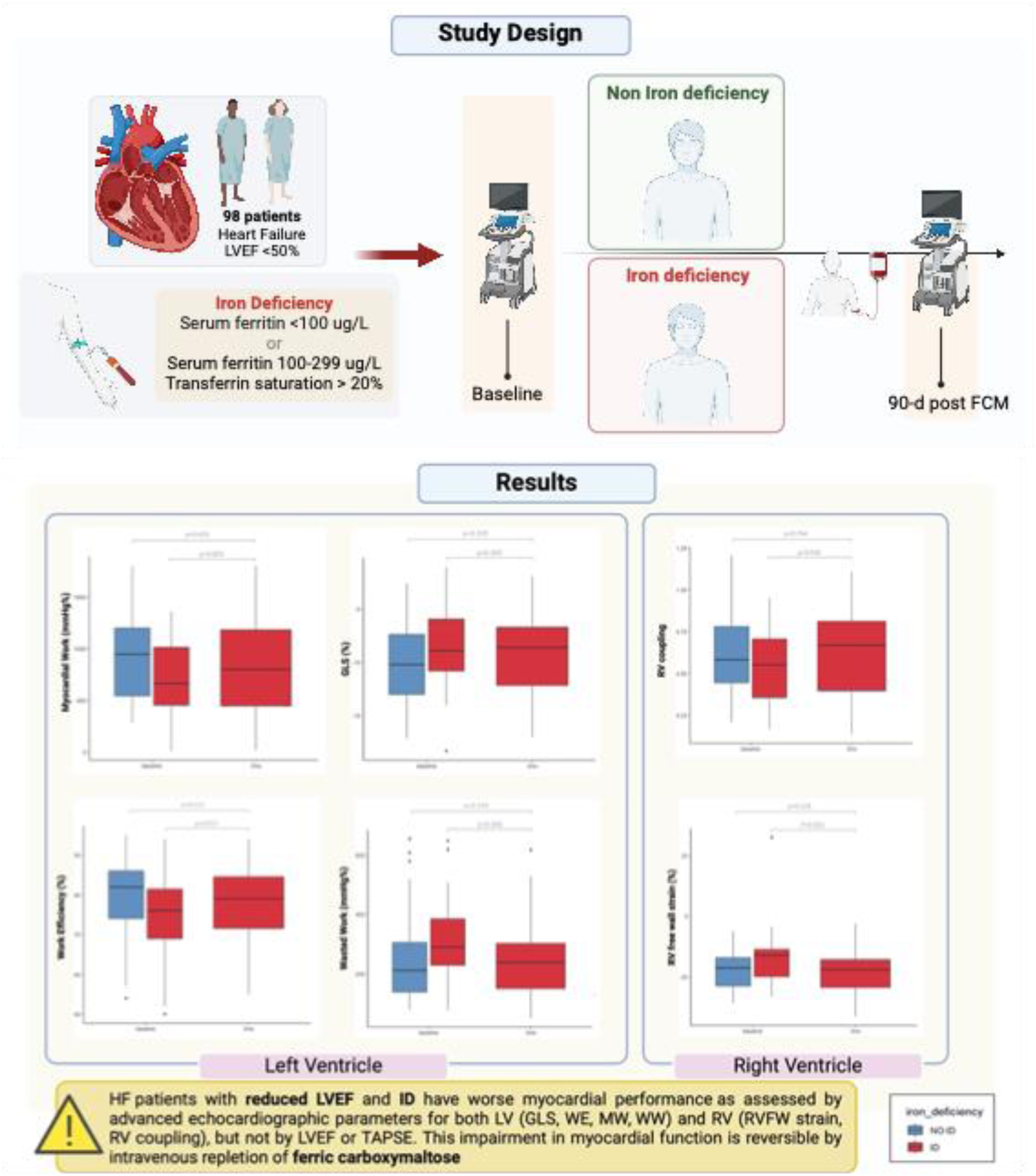

## Introduction

Iron deficiency (ID) is highly prevalent among heart failure (HF) patients (1). Regardless of the presence of anemia, ID reduces the energy efficiency of nonhematopoietic tissues with high energy demands (2). Thus, ID affects the function of the cardiomyocytes by impairing mitochondrial respiration chain, ATP production and contractility (3). The decline in myocardial function due to ID has a pathophysiological basis, which has been explored both in human and animal models (4). Therefore, anemia and ID directly affect cardiac function since they lead to a decrease in preload and the size of the left ventricular cavity (5).

Iron repletion in patients with HF and left ventricular ejection fraction (LVEF) of less than 50% is associated with improvements in functional capacity, quality of life and reduction of rehospitalization (6–10).However, the mechanisms by which iron replenishment improves exercise capacity and quality of life remain unclear.

Although previous studies have shown that iron therapy may be linked to enhancements in cardiac function (5,11), the surrogate parameters of myocardial function used in these studies have several limitations. First, LVEF is a volume-derived index, relies on geometric assumptions, and is load-dependent (12). Second, in spite of speckle-tracking-derived longitudinal strain offers lower inter-and intra-observer variability, conditions of elevated pre-or after-load directly affect GLS (13). Regarding this, the development of new echocardiography software has provided access to non-invasive assessments of highly valuable and more accurate information on cardiac physiology via myocardial work (MW) index and its derivates constructive work (CW) and wasted work (WW), directly obtained by measuring the area of the pressure-strain loop (14). Finally, noninvasive assessment of ventricular-arterial coupling (VAC) may be useful for a more complete cardiovascular performance profiling in this setting.

Interestingly, none of the previous studies that have attempted to assess the impact of iron status on cardiac function parameters have explored the impact of iron deficiency on myocardial performance parameters. This information is crucial to explain the mechanisms that drive the worse exercise capacity observed in patients with ID (15). Finally, the reversibility of the potential myocardial performance alterations associated with ID after iron replacement is unknown.

Given the gaps of knowledge in this field and the limitations of previous studies, the IRON-PATH II echo-substudy was designed to define cardiac remodeling and myocardial deformation associated with systemic and tissue ID in patients with HF and reduced LVEF, compared to patients with HF without ID. Secondly, we aim to explore changes in myocardial function after iron replacement using echocardiography.

## Methods

### Study design

The IRON-PATH II (New pathophysiological pathways involved in iron metabolism disorder in hear failure: The IRON PATH II investigator-initiated Study, NCT05000853) was a multicenter, prospective, observational, and investigator-initiated study, aimed to characterize the biological pathways involved in ID in HF patients and its association with laboratory biomarkers and clinical outcomes. The IRON-PATH II echo-substudy was designed to describe the echocardiographic features of HF patients according to ID and the effects of intravenous ferric carboxymaltose repletion on echocardiographic parameters.

The IRON PATH II study included a total of 210 HF patients (80 patients without ID and 130 patients with ID) recruited in 7 centers across Spain and Portugal between August 2021 and May 2023 and followed for a fixed period of 12 months. The echo-substudy included a subset of 100 patients, all recruited in the coordinating center, in whom a specific echocardiographic evaluation was performed.

The IRON-PATH II study was sponsored by CSL Vifor with coordination by Bellvitge Biomedical Research Institute (IDIBELL). The sponsor didńt participated in the design of the study nor data analysis. An independent data and safety monitoring committee reviewed data every 6 months. The study was approved by the local ethics committees for clinical research at each participating center and was conducted in accordance with the principles of the Declaration of Helsinki. All patients gave written informed consent before study entry. The echo-substudy was included as a specific analysis within the IRON-PATH II project for those patients included in Bellvitge University Hospital. Safety was reported by local investigators in accordance with the current legislation that regulates pharmacovigilance in Spain and Portugal. End-point adjudication for the components of the primary outcome of the study has been performed by an independent end-point committee blinded to the group allocation of patients on an ongoing basis according to the reporting of major events (death, readmission) made by the local investigators.

### Patients

Patients with HF and LVEF ≤50% according to the European Society of Cardiology (ESC) diagnostic criteria (16), receiving oral medication for chronic HF according to GDMT and without signs of fluid overload or low cardiac output were included. Reasons for ineligibility included significant anemia (Hb <11 g/dL), undergoing erythropoiesis-stimulation agents, oral iron supplementation or intravenous iron treatment within the last 3 months. Individuals with a planned resynchronization therapy, coronary revascularization, heart transplant or left ventricular assist device implantation or with less than one year of life expectancy were also excluded.

### Study Procedures

Patients were classified according to the current ESC guidelines definition of ID (serum ferritin <100ug/l or ferritin 100 – 299ug/l with TSAT <20%)(16). ID patents were treated following clinical recommendations; dosage was calculated according to their weight and hemoglobin levels. Ferric carboxymaltose (FCM, Ferinject, CSL Vifor) was administered as intravenous perfusion diluted in sterile solution administered over 30-60 minutes. The patients received a maximum of 1000 mg of iron at once. In those patients in whom more iron was needed, a second dose was administered, at least fifteen days apart from the first one.

### Study data

A detailed baseline evaluation was performed for all participants at study entry. Demographic characteristics, clinical and disease related factors, co-morbidities, laboratory tests and medical treatments were collected. Patients with ID were re-assessed after 3 months of iron replenishment (clinical evaluation, blood test and echocardiography was performed). Clinical events were evaluated in an ongoing basis in all patients and finally ascertained after 12 months of follow-up.

### Echocardiographic evaluation

Echocardiographic assessment was performed at baseline. In the ID group, echocardiography was repeated 3 months after intravenous iron replacement. Echocardiographic examinations were done by two experienced research sonographers blinded to the iron status of the patient and the therapeutic interventions conducted in terms of iron repletion. Standard commercial Vivid E97 equipment and a MC5 active-matrix transducer (GE Medical Systems) was used. All the images were digitally stored online and measured with specific offline software (EchoPAC Version 202, GE Medical Systems). All images were acquired throughout a complete cycle at a frame rate that will represent 80% of the heart rate without exceeding 90 Hz or falling below 40 Hz. The data were transferred for further analysis in specific software. Measurements were made in three cycles (five cycles in those patients in atrial fibrillation during the examination) and the average value was calculated.

Echocardiographic features included bidimensional and diastolic parameters (septal wall, posterior wall, LV end-diastolic diameter, LV end-diastolic volume, LA indexed volume, E/A ratio, E/e’ ratio), left and right ventricle function parameters (LVEF, GLS, LV indexed stroke volume, LV indexed stroke work, cardiac output, TAPSE, systolic pulmonary artery pressure, FAC, RV coupling ratio, RV free wall strain) and vascular function parameters (systemic arterial compliance, systemic arterial resistance index, LV end-systolic elastance (Ees), arterial elastance (Ea), ventricular-arterial coupling) (**Table S1)**.

MW was assessed by the combination of LV strain data, the non-invasively estimated LV pressure curve (brachial cuff blood pressure) and the valvular event timing (14). Strain and pressure data were synchronized using the onset of R-wave at EKG, and the area of the pressure strain loop is used to derive segmental and global MW. Global work index (GWI) was calculated as the average of segmental values. CW was defined as work during segmental shortening in systole and during lengthening in isovolumic relaxation. WW (energy loss) was the work performed during lengthening in systole and shortening in isovolumic relaxation. The work efficacy (WE) was automatically calculated as the ratio of CW/(CW+WW).

### Study outcomes

The main endpoint was the change in cardiac performance, including LVEF and LV deformation parameters (GLS and its derivates MW, WW, CW and WE) after iron replenishment. Secondary endpoints included change in RV function parameters (TAPSE, FAC, RV free wall strain and RV coupling ratio). Finally, exploratory endpoints included cardiovascular function explored by Ea, Ees and VAC.

## Statistical analysis

Variables are expressed as mean and standard deviation (in case of normal distribution) or median and interquartile range (in case of non-normal distribution). Boxplots were used to graphically describe the distribution of the variables. Comparison was performed using chisquare (for qualitative variables), t-test (for quantitative variables with normal distribution) or U-Mann-Whitney-Wilcoxon (for quantitative variables with non-normal distribution). The t-test was used for those variables that met the assumptions of independence, normality (using the Kolmogorov-Smirnov test) and homoscedasticity or equality of variances (using the Levene test).

The comparison between baseline echocardiographic parameters (before iron replenishment) and those after 3 months since the iron replenishment was performed using a t-test for paired data. The 3-month variables of the iron-deficient group (after iron replenishment) were compared with the baseline variables of the non-iron-deficient group. Bar graphs were used to graphically describe the evolution of the variables. No multivariate statistical model was performed.

All statistical tests and confidence intervals (CI) were constructed with a Type I error, alpha level of 5%, with no adjustments for multiplicity. P values below 0.05 were considered statistically significant. All analyses were performed using the SPSS software (version 22.0; IBM, Armonk, NY) and R software packages (version 4.2.1; R Foundation for Statistical Computing, Vienna, Austria).

## Results

The IRON-PATH II echo-substudy included a subset of 100 patients undergoing a specific echocardiographic evaluation. Two patients were excluded due to a poor sonographic window; 98 patients were finally analyzed (**Figure S1**).

### Baseline Characteristics

Baseline characteristics of the whole IRON-PATH II study cohort are presented in **Table S2**. Patients included in the echo-substudy cohort were slightly older (72 vs 68 years, p=0.027) and with worse renal function (eGFR 55 vs 60 ml/min/1.73m2, p=0.044). Ferritin levels were lower in patients not included in the echo-substudy cohort (ferritin 244 vs 138 ng/mL, p<0.001) although no differences in TSAT were observed (22 vs 21%, p=0.351).

The baseline characteristics of the echo-substudy sample, both overall and according to ID status, are listed in **Table 1**. ID was present in 44 patients (45%). Considering the clinical profile, ID patients were older (74±8 vs 70±11 years; p-value = 0.03), more frequently male (78%), NYHA class II (71%) and had slightly higher heart rate (72 vs. 67 bpm, p=0.043). They also covered shorter distances in the 6-minute walking distance (206 vs 314 m, p<0.001). Comorbidities appeared to be similar in the two groups, with the exception of atrial fibrillation (28 vs 22%, p-value = 0.042).

**Table 1.**
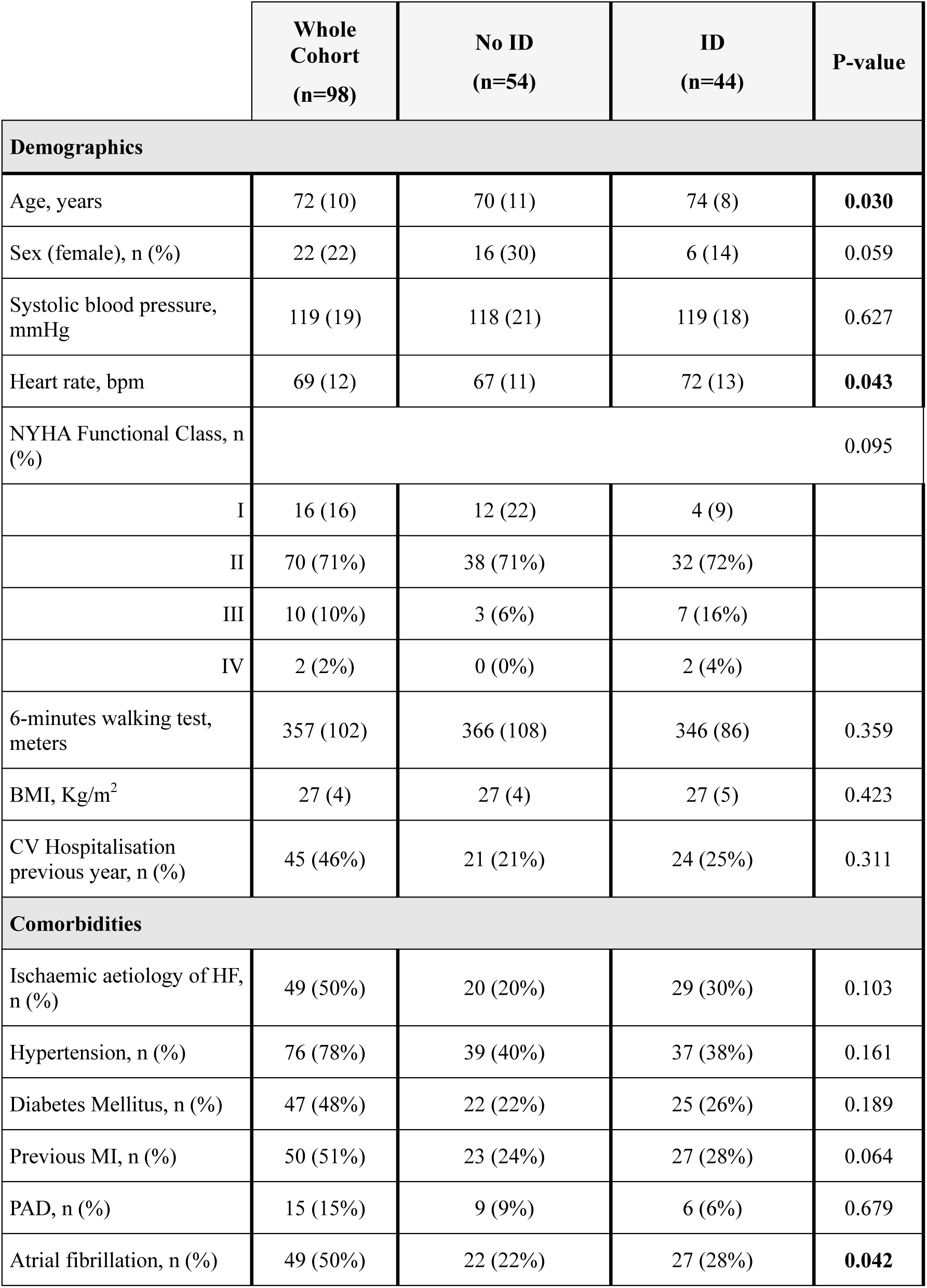
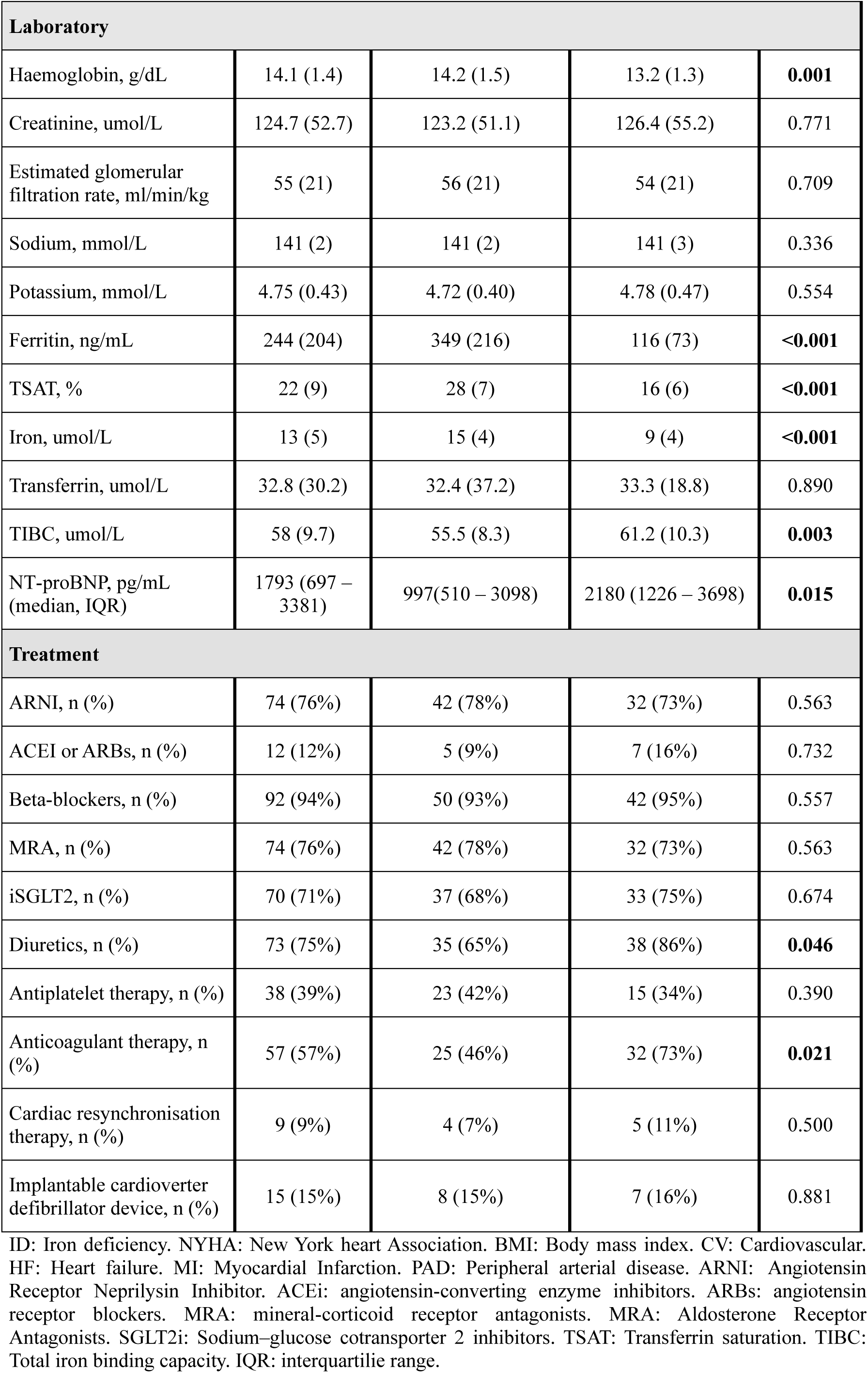
Demographic and clinical characteristics of all patients included in this analysis, overall and according to iron status.

The mean hemoglobin levels were lower among ID patients (13.2 vs. 14.2, p = 0.001) and the NTproBNP was higher (2180 vs 997, p-value = 0.015). ID group showed marked iron depletion evidenced by lower transferrin saturation (TSAT) (16 vs 28%), ferritin (116 vs 349 ng/mL) and iron (9 vs 15 umol/L) (all p-value <0.001). Both groups were similarly treated (**Table 1**) with beta-blockers, angiotensin receptor neprilysin inhibitors (ARNI), sodium-glucose cotransporter-2 (SGLT2i) and mineralocorticoid receptor antagonist (MRA) (94%, 76%, 71% and 76%, respectively), all p-values >0.05. Anticoagulation therapy was more frequent among ID patients (46% vs 73%, p-value=0.021).

### Echocardiographic features at baseline

The groups according to ID status showed no differences concerning LV thickness, LV volumes, E/A ratio or E/e’ ratio (all p-values >0.05) (**Table 2**). No differences were observed in LVEF (38±10% vs 35±9%, p-value=0.210), TAPSE (18 mm vs 17 mm, p-value=0.118) or sPAP (29 mmHg vs 30 mmHg, p-value=0.150). ID group displayed worse LV function expressed by lower GLS (–8.5±9% vs –10±10%, p-value=0.024), lower work efficiency (74±10% vs 80±10%, p-value=0.017), lower myocardial work (665[453-1013] mmHg% vs 947[542-1199] mmHg%, p-value=0.025) and higher wasted work (290[228-384] mmHg% vs 212[138-305] mmHg%, p-value=0.034) (**Table 3 and Figure S2, panel A**). RV function was also worse as shown by lower RV free wall strain (–13[-20-(– 11)] % vs –17[-23-(–14)] %, p-value=0.022) (**Table 3 and Figure S2, panel B**). However, no differences were observed regarding TAPSE (18 vs 17 mm, p-value=0.118) or sPAP (29[24-35] vs 30[26-42] mmHg, p-value=0.15). Finally, the ID group obtained worse end-diastolic elastance values (3.0 vs 2.1 mmHg/mL, p-value=0.03), and a worse ventricle-arterial coupling ratio (0.95 vs 1.4 mmHg/mL, p-value = 0.04) (**Table 3 and Figure S2, panel C**).

**Table 2.** Echocardiographic characteristics of all patients included in this analysis, overall and according to tissue iron status.

|  | Whole Cohort<br>(n=98) | No ID<br>(n=54) | ID<br>(n=44) | P-value |
| --- | --- | --- | --- | --- |
| Septal wall, mm | 11 (3) | 11 (3) | 11 (3) | 0.890 |
| Posterior wall, mm | 10 (2) | 10 (2) | 10 (2) | 0.267 |
| LV end-diastolic diameter, mm | 55 (10) | 55 (10) | 55 (10) | 0.803 |
| LV indexed end-diastolic volume, mL/m<sup>2</sup> | 78 (31) | 76 (32) | 80 (28) | 0.452 |
| LV indexed end-systolic volume, mL/m<sup>2</sup> | 50 (26) | 46 (27) | 54 (25) | 0.182 |
| LA indexed volume, mL/m<sup>2</sup> | 49 (17) | 45 (17) | 53 (16) | **0.026** |
| E/A ratio | 1.4 (1) | 1.3 (1) | 1.5 (1) | 0.661 |
| E/e' ratio | 13 (7) | 13 (6) | 14 (9) | 0.596 |
| LVEF, % | 36 (9.66) | 38 (10.26) | 35 (8.78) | 0.210 |
| TAPSE, mm | 17 (4.18) | 18 (4.24) | 17 (4.03) | 0.118 |
| Systolic pulmonary artery pressure, mmHg (median, IQR) | 30 (25 – 37) | 29 (24 – 35) | 30 (26 – 42) | 0.15 |

LV: Left ventricle. LVEF: Left ventricle ejection fraction. GLS: Global longitudinal strain. TAPSE: Tricuspid Annular Plane Systolic Excursion.

**Table 3.** T-test and U-Mann-Withney exploring echocardiographic characteristics differences before (ID baseline) and after (ID 3rd month) intravenous iron replacement (in the ID group), as well as after iron replacement (ID 3rd month) versus the non-ID group.

|  | **Whole Cohort**<br>**(n=98)** | **No-ID** | **ID baseline**<br>***before iron***<br>***replacement*** | **ID 3<sup>rd</sup> month**<br>***after iron***<br>***replacement*** | **P-value**<br>***ID baseline***<br>***vs no-ID***<br>***baseline*** | **P-value**<br>***ID baseline***<br>***vs ID 3<sup>rd</sup>***<br>***month*** | **P-value**<br>***ID 3<sup>rd</sup> month***<br>***vs no-ID*** |
| --- | --- | --- | --- | --- | --- | --- | --- |
| ***Left ventricular function*** |  |  |  |  |  |  |  |
| LVEF, % | 36 (9.66) | 38 (10.26) | 35 (8.78) | 36 (9.66) | 0.210 | 0.06 | 0.203 |
| GLS, % | -9.5 (3.69) | -10.2 (10.26) | -8.5 (8.78) | -9.31 (3.43) | **0.024** | **0.003** | 0.329 |
| LV indexed stroke work, g.m/m<sup>2</sup> | 33 (13) | 34 (9) | 32 (17) | 31 (10) | 0.423 | 0.422 | 0.564 |
| Cardiac output, L/min (median, IQR) | 4.7 (3.8 – 5.7) | 4.7 (3.78 – 5.90) | 4.61 (3.93 – 5.46) | 4.2 (3.50 – 4.40) | 0.62 | **0.08** | 0.131 |
| Cardiac index, L/min/m<sup>2</sup> (median, IQR) | 2.4 (2 – 2.5) | 2.6 (2.1 – 3) | 2.7 (2.5 – 2.9) | 2.3 (2 – 2.7) | 0.728 | **0.075** | 0.135 |
| Myocardial work, mmHg% (median, IQR) | 823 (504 – 1113) | 947 (542 – 1199) | 665 (453 – 1013) | 801 (447 – 1183) | **0.025** | **0.003** | 0.435 |
| Constructive work, mmHg% | 1131 (449.10) | 1191 (449.82) | 1053 (441.63) | 1108(471) | 0.149 | 0.06 | 0.420 |
| Wasted work, mmHg% (median, IQR) | 262 (155 – 361) | 212 (138 – 305) | 290 (228 – 384) | 239 (151 – 302) | **0.034** | **0.006** | 0.919 |
| Work efficiency, % | 77.41 (10.37) | 80 (9.93) | 74(10.32) | 79 (9.98) | **0.017** | **0.017** | 0.521 |
| *Right ventricular function* |  |  |  |  |  |  |  |
| TAPSE, mm | 17 (4.18) | 18 (4.24) | 17 (4.03) | 17 (4.31) | 0.118 | 0.118 | 0.989 |
| Systolic pulmonary artery pressure, mmHg (median, IQR) | 30 (25 – 37) | 29 (24 – 35) | 30 (26 – 42) | 28.10 (24.72 – 40.28) | 0.15 | 0.58 | 0.73 |
| FAC, % (median, IQR) | 45 (38 – 52) | 45.45 (38.74 – 54.69) | 45.19 (36.93 – 49.66) | 44 (35.45 – 47.57) | 0.095 | 0.68 | 0.162 |
| RV coupling ratio, mm/mmHg | 0.59 (0.23) | 0.63 (0.25) | 0.54 (0.21) | 0.61 (0.27) | 0.083 | **0.036** | 0.764 |
| RV free wall strain, % (median, IQR) | -16 (-22 – -12) | -17 (-23 – -14) | -13 (-20 – -11) | -17.6 (-23.5 – -14.3) | **0.022** | **<0.001** | 0.528 |
| *Cardiovascular function* |  |  |  |  |  |  |  |
| Systemic arterial compliance, mL/m<sup>2</sup>/mmHg (median, IQR) | 1.3 (1 – 1.6) | 1.26 (1.01 – 1.56) | 1.27 (1.04 – 1.63) | 1.1 (0.90 – 1.51) | 0.93 | 0.16 | 0.09 |
| Systemic arterial resistance index, dyn*s*cm<sup>-5</sup>.m<sup>-2</sup> | 2449 (764.39) | 2442 (751) | 2458 (789) | 2690 (822) | 0.923 | 0.10 | 0.217 |
| LV end-systolic elastance, mmHg/mL (median, IQR) | 2.5 (1.7 – 3.5) | 3.0 (1.9 – 3.6) | 2.1 (1.5 – 3.1) | 2.34 (1.53 – 3.42) | **0.03** | 0.20 | 0.511 |
| Arterial elastance, mmHg/mL (median, IQR) | 2.8 (2.4 – 3.4) | 2.7 (2.4 – 3.3) | 3.0 (2.4 – 3.7) | 3.26 (2.73 – 3.80) | 0.26 | 0.1 | 0.011 |
| Ventricular-arterial coupling (ratio) (median, IQR) | 1.1 (0.8 – 1.9) | 0.95 (0.75 – 1.4) | 1.4 (0.9 – 2.2) | 1.46 (0.86 – 2.37) | **0.04** | 0.53 | 0.109 |

LV: Left ventricle. LVEF: Left ventricle ejection fraction. GLS: Global longitudinal strain. TAPSE: Tricuspid Annular Plane Systolic Excursion. FAC: Fractional Area Change. RV: Right ventricle

### Echocardiographic features after iron repletion

After intravenous iron replacement (**Table 3**), improvements in GLS (–8.5±9% vs –9.3±3%), work efficiency (74±10% vs 79±10%), myocardial work (665[453-1013] mmHg% vs 801[447-1183] mmHg%) and wasted work (290[228-384] mmHg% vs 239[151-302] mmHg%) were observed (all p-values <0.05). Attending to RV function, patients improved RV coupling ratio (0.54±2 vs 0.61±0.3 mm/mmHg, p-value=0.036) and RV free wall strain (–13 [-20 to –11]% vs –18 [-24 to –14]%, p-value <0.001). Iron replacement had no impact on LV end-diastolic elastance or ventricle-arterial coupling (Ees/Ea). No differences were observed in any echocardiographic parameter of myocardial function between non-ID patients in comparison with ID-patients after iron replenishment (**Figure 2 and Table 3**).

## Discussion

Our study provides relevant information regarding the pathophysiology of ID in the HF and LVEF <50% scenario. Firstly, patients with HF and ID displayed worse myocardial performance than non-ID patients, both for LV (GLS, myocardial work, wasted work and work efficiency) and RV (RV free wall strain and RV coupling), with no differences in LVEF or TAPSE. Secondly, iron repletion significantly improved myocardial performance in ID patients, resulting in myocardial performance phenotype similar to non-ID patients (**Graphical Abstract**).

There is accumulating evidence that ID is not only a consequence but also a contributor to the pathophysiology of systolic HF, potentially exacerbating disease progression through mitochondrial dysfunction caused by intracellular iron depletion (17–19). Previous studies showed that correcting anemia and ID results in improvement in echocardiographic parameters, suggesting the role of these conditions in myocardial mechanics (5). Toblli et al. demonstrated enhancement in LVEF (6.6 ±3.8%) and reduction in LV diameters (LVESD 10.5±1.6 mm, LVEDD 9±13 mm) with iron sucrose therapy, among HF with reduced LVEF patients (20). The IRON-CRT study (21) included 75 patients with HF, ID criteria and LVEF of 45% or less. After CRT therapy, a greater increase in LVEF in response to CRT was observed in the group in which FCM was replaced compared to placebo. Response was analyzed 3 months after iron replacement. The improvement was due to LV end-systolic volume (LVESV) reduction, with no differences in LV end-diastolic volume (LVEDV).

Scarce evidence exists regarding the effect of iron repletion on strain parameters (11). To our knowledge, only the Myocardial-IRON substudy (22) has explored the effect of iron repletion on myocardial function expressed through strain parameters. Improvement in global longitudinal, circumferential and radial strain of both LV and RV was observed 30 days after iron repletion. In COPD patients without anemia, ID was associated with a moderate increase in systolic pulmonary artery pressure (23). However, no work to date has assessed right ventricular remodeling associated with iron deficiency. Thus, in patients with HF the impact of iron replacement on RV has not been adequately explored.

### Myocardial work as a surrogate of cardiac function, back to the pressure-volume loop

Our cohorts showed no differences in LVEF, LV stroke volume, LV stroke work, TAPSE or sPAP (all p values >0.05). Nevertheless, in the IRON-PATH II echo-substudy we employed speckle-tracking echocardiography (GLS, myocardial work index and its derivates) and VAC assessment as myocardial performance subrogates. These tools offer significant advantages over traditional measures like LVEF, as they offer a more accurate assessment of myocardial function by integrating both cardiac and arterial components (13,24,25). The analysis of pressure-volume loops allows the measurement of the energy imparted to the blood, with the area of the loop representing stroke work (SW)(26). New echocardiography software creates arterial pressure–LV longitudinal myocardial strain curves by speckle tracking (14). This software calculates myocardial work, constructive work (work during segmental shortening in systole and during lengthening in isovolumic relaxation) and waste work (lengthening in systole and shortening in isovolumic relaxation).

Non-invasive methods for assessing myocardial work have shown good agreement with invasive pressure-volume loop measurements (27). Thus, myocardial work index allows for a comprehensive assessment of the energy dynamics within the heart taking into account loading conditions. In addition to myocardial work index (work evaluated from mitral valve closure valve opening), waste work is calculated as myocardial work consumed during segmental lengthening (negative work) that does not turn into segmental contraction (positive work)(14,27).

These parameters have been previously explored in the HF scenario. Sacubitril-Valsartan treatment has been shown to improve constructive work and work efficiency (28); likewise, these two parameters have been proposed as a predictor of events in advanced HF (29) and as a predictor index of CRT-response (30). To the best of our knowledge, this is the first study demonstrating myocardial work improvement after iron repletion, due to a decrease in waste work and an improvement in work efficiency. RV functional parameters as RV coupling ratio and RV free wall strain also improved. Interestingly, after iron repletion these functional parameters were similar to non-ID group.

Beyond myocardial performance, VAC permits to study the relationship between the myocardium and the arterial tree. VAC is evaluated by non-invasively measuring the ratio of arterial (Ea) to ventricular end-systolic elastance (Ees) (31)(32). Ea is an integrative measure of arterial system properties (33), while Ees defines the ventricle’s contractile state and is relatively insensitive to loading conditions. Thus, VAC evaluation together with the interpretation of its components (Ea and Ees) allows for a more complete myocardial performance profile in patients with heart failure (HF) and iron deficiency (ID).

Ventricle-arterial uncoupling in HFrEF is characterized by a decrease in Ees together with an increase in Ea (derived from neurohormonal activation), which leads to higher VAC values (31). Either CRT implantation (34) and pharmacological treatment with β-blocker (35) may improve VAC. Patients with ID showed worse VAC at the expense of lower Ees. However, no differences in Ees, Ea or VAC were observed after iron replacement.

### The role of iron in myocardial performance

Myocardial activity is closely linked to oxidative metabolism (36). Notably, not all the energy generated by oxidative metabolism is utilized and converted into effective work. Therefore, our data suggests ID may contribute to HF progression based on the lower myocardial work (likely related with the higher waste work displayed by ID patients), and the reversibility after iron replacement.

Clinically, the iron-deficient group was probably composed of patients with a true iron-deficient state, as reflected by a mean TSAT of 16%. The significant improvements observed in our study can be explained by the restoration of mitochondrial function and energy production in cardiomyocytes due to iron repletion (37). Notably, ID repletion had positive effect on cardiac performance even that AF was more frequent and mean left atria was larger in ID group in patients. Finally, the positive changes in RV function and ventricular-arterial coupling suggest that iron therapy may also improve pulmonary hemodynamics and right ventricular performance, contributing to better overall heart function.

Our study highlights the significant improvements in myocardial performance following intravenous iron repletion in patients with HF and ID. These findings underscore the potential of iron therapy to enhance cardiac function beyond traditional measures such as LVEF or TAPSE. Future research should include mechanistic studies to elucidate the pathways through which medical therapies in general and iron repletion in particular, benefit myocardial function. These insights will pave the way for more targeted and effective therapeutic strategies.

## Limitations

Our study has some limitations that should be mentioned. First, our study is based on echocardiography, so the performance of cMRI in the study of myocardial work after iron replacement is uncertain. However, echocardiography is an accessible technique and myocardial work calculation is an easy-use and reproducible tool, as it has been included in echocardiographic software. Second, speckle-tracking requires experience and depends on image quality. Noninvasive arterial pressure is included in the calculation of myocardial work, which can be variable throughout the cardiac cycle. Third, both groups were well balanced. However, ID patients were slightly older, had higher prevalence of AF and lower mean hemoglobin levels. Although this may partially explain the baseline differences, the improvement in myocardial performance after iron replacement was clear. Finally, both groups underwent baseline echocardiography but only the ID group received control echocardiography (3 months after replacement). Thus, direct comparison between the two groups is limited.

## Conclusions

The IRON-PATH II echo substudy demonstrates that HF patients with reduced LVEF and ID have worse myocardial performance of both the LV and RV. Specifically, ID patients had impaired GLS, work efficiency, myocardial work, waste work, RV free wall strain, and RV coupling. No differences were observed on LVEF or TAPSE. These alterations in myocardial function were measurable by echocardiography and were reversible by intravenous repletion of ferric carboxymaltose. This underscores the significant role of iron therapy in enhancing cardiac function. Future research should explore the underlying mechanisms and further validate these findings to optimize therapeutic strategies.

## Data Availability

All data supporting the findings of this study are available upon reasonable request from the corresponding author. De-identified participant data can be shared in accordance with institutional and ethical guidelines.

## Abbreviations

AF: Atrial fibrillation
ARNI: Angiotensin receptor neprilysin inhibitor
ATP: Adenosine triphosphate
CRT: Cardiac resynchronization therapy
CW: Constructive work
Ea: Arterial elastance
Ees: End-systolic elastance
ESC: European Society of Cardiology
FCM: Ferric carboxymaltose
FAC: Fractional area change
GLS: Global longitudinal strain
GWI: Global work index
Hb: Hemoglobin
HF: Heart failure
ID: Iron deficiency
LVEF: Left ventricular ejection fraction
LVEDV: Left ventricular end-diastolic volume
LVESD: Left ventricular end-systolic diameter
LVESV: Left ventricular end-systolic volume
MW: Myocardial work
MRA: Mineralocorticoid receptor antagonist
NTproBNP: N-terminal pro-brain natriuretic peptide
NYHA: New York Heart Association
RV: Right ventricle
RVFW: Right ventricular free wall
sPAP: Systolic pulmonary artery pressure
TAPSE: Tricuspid annular plane systolic excursion
TSAT: Transferrin saturation
VAC: Ventricular-arterial coupling
WE: Work efficiency
WW: Wasted work

## Acknowledgments

The authors kindly acknowledge all study participants in the IRON-PATH II study. We thank CERCA Programme/Generalitat de Catalunya for their institutional support. We thank Departament de Salut de la Generalitat de Catalunya through “Pla Estratègic de Recerca i Innovació en Salut (PERIS 2023)” grant number SLT028/23/000018 for financial support.

---

**Figure 1.**
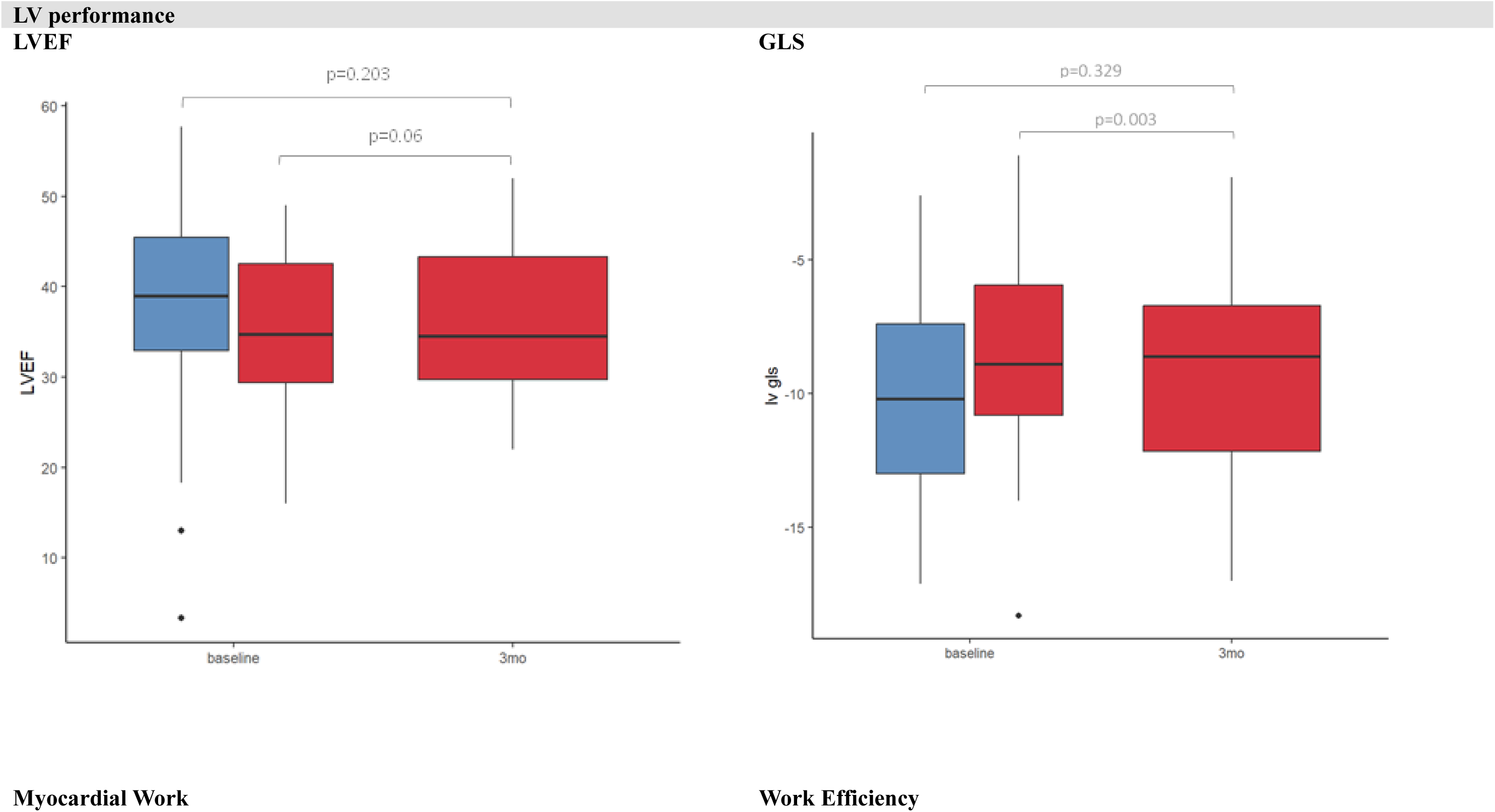
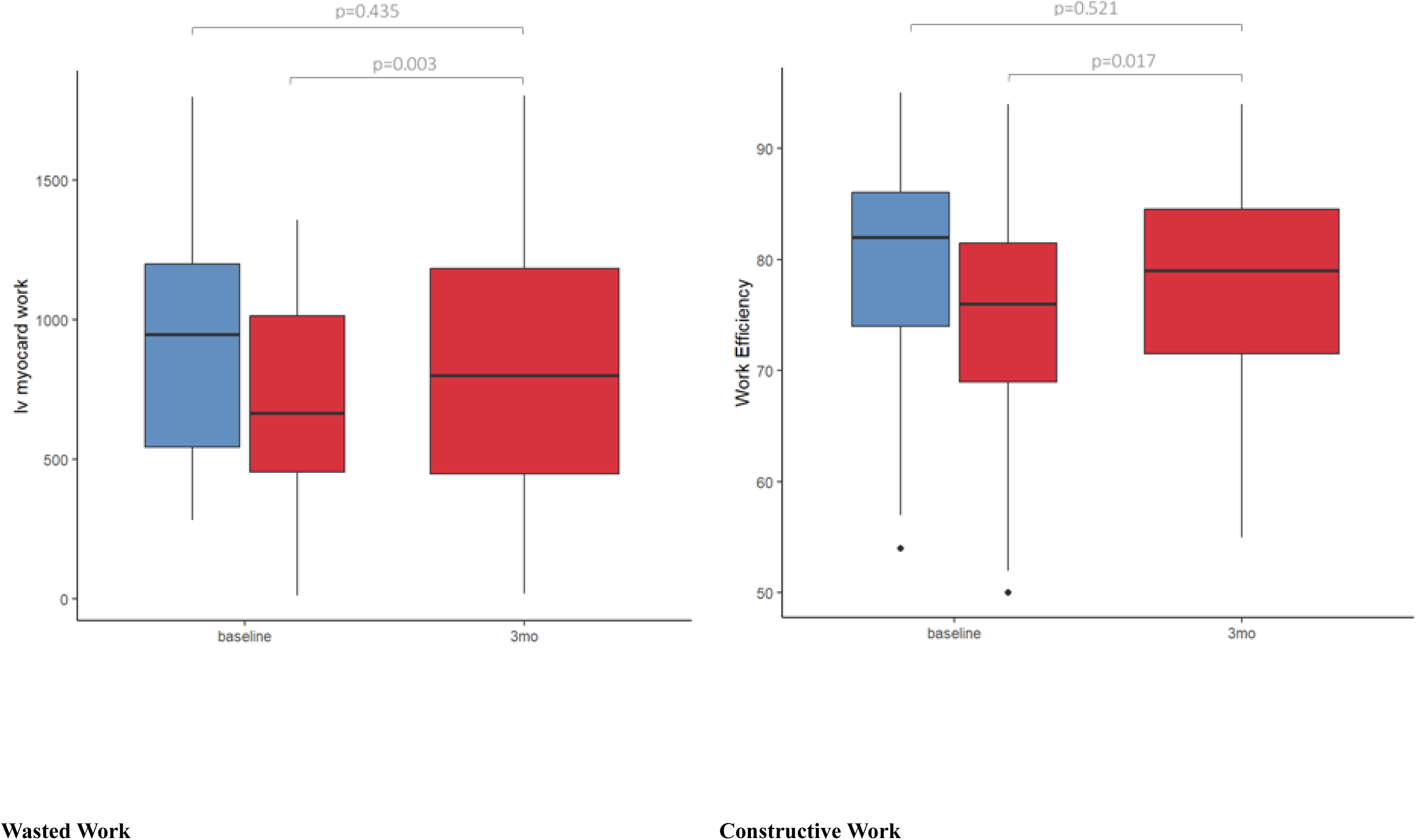
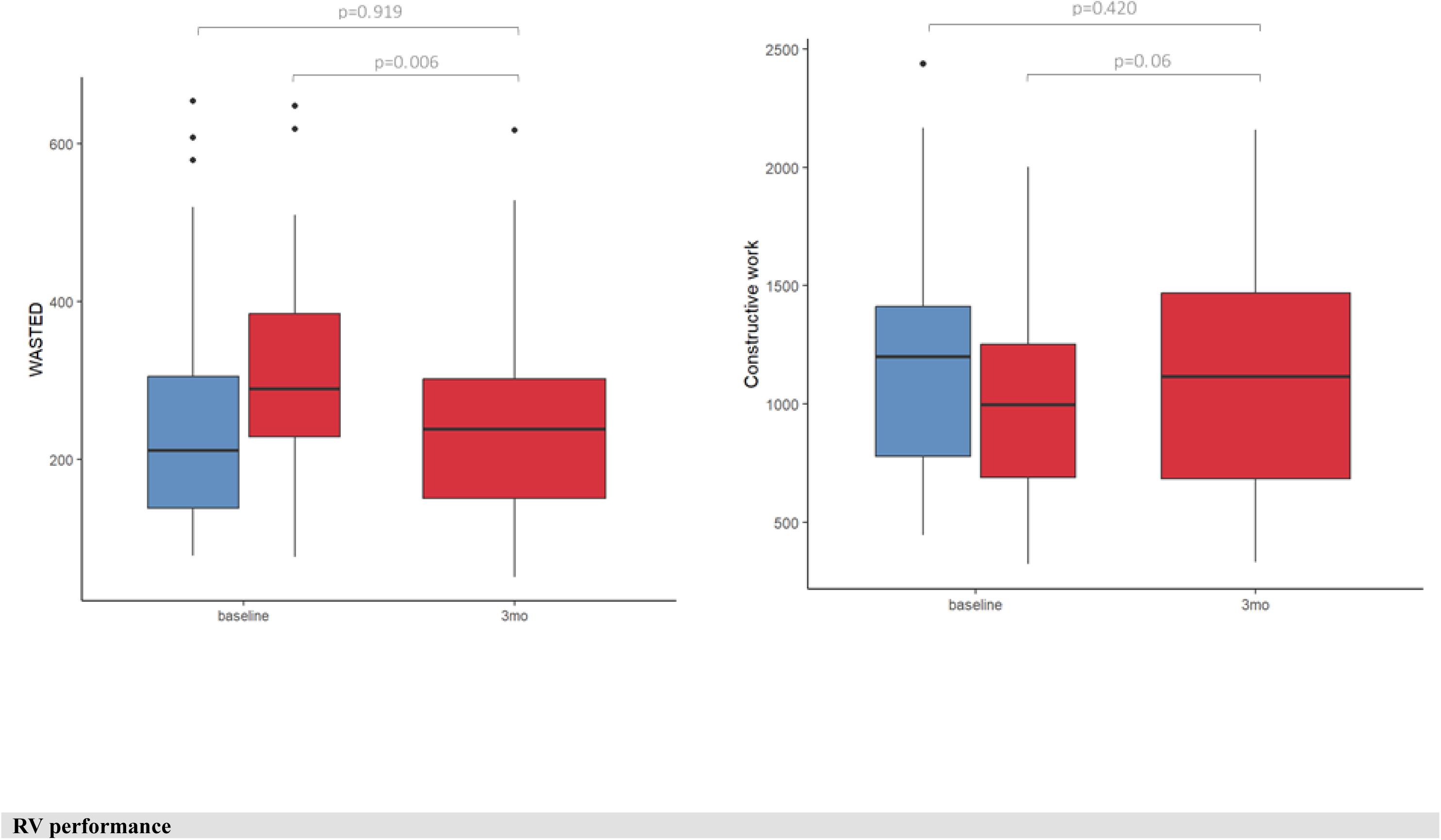
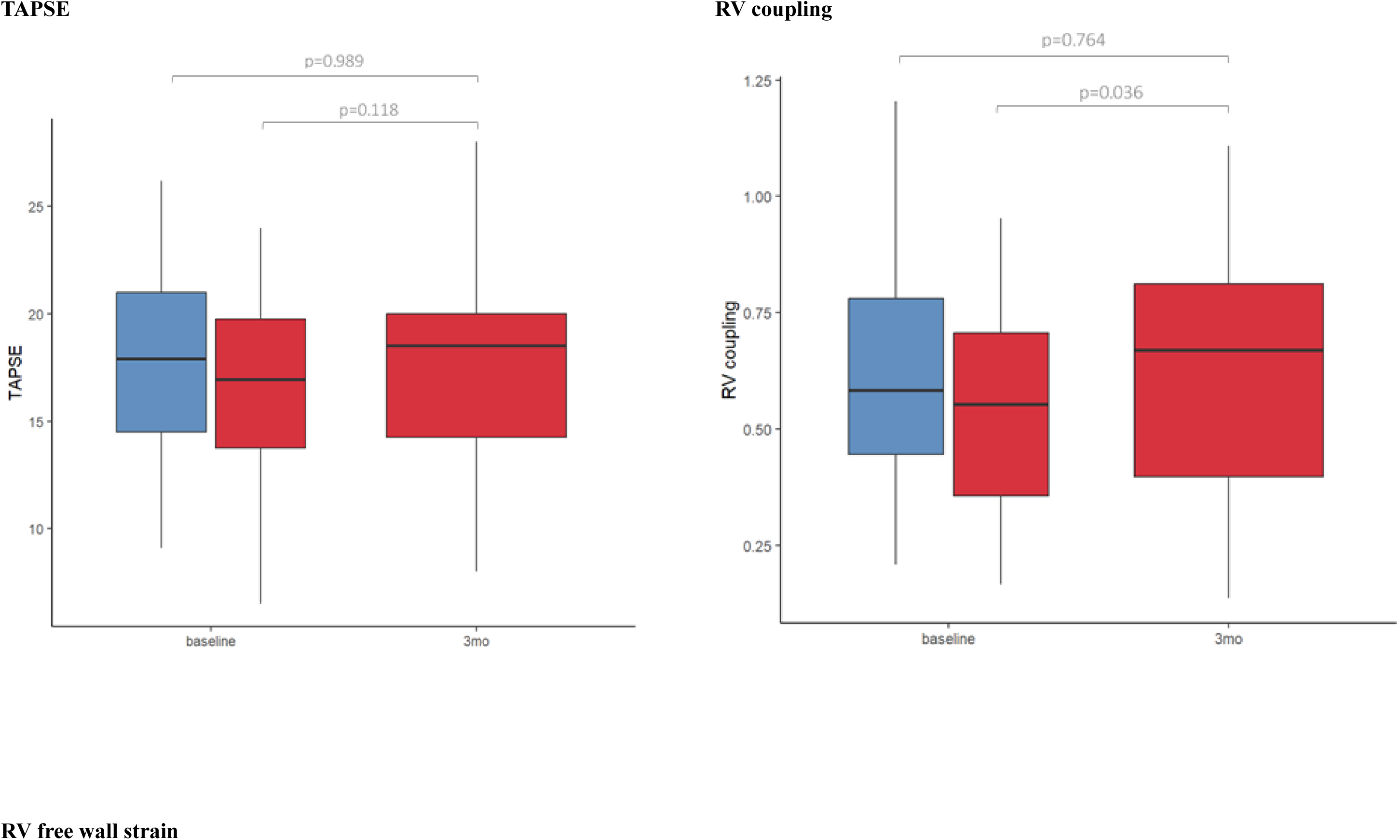
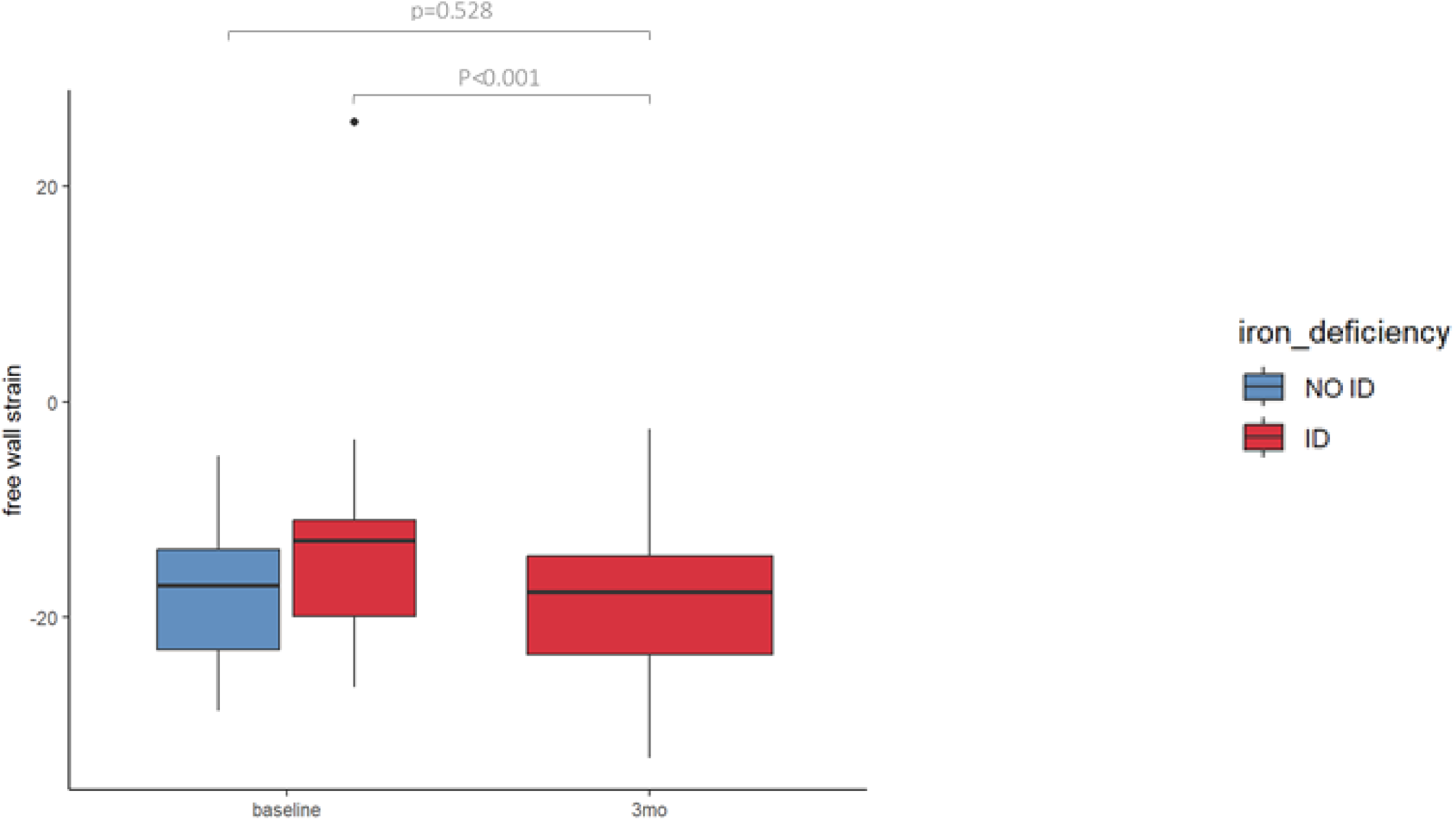
Bar graphs (showing mean and standard deviation) of LV myocardial performance (LVEF, GLS, myocardial work, constructive work, wasted work and work efficiency) and RV myocardial performance (TAPSE, RV coupling, FAC, RV free wall strain). No ID (blue boxes) vs ID (red boxes).

